# Modelling the Occurrence of the Novel Pandemic COVID-19 Outbreak– A Box and Jenkins Approach

**DOI:** 10.1101/2020.06.15.20131136

**Authors:** Ajadi Nurudeen, Ogunsola Isqeel Adesegun, Damisa Adams Saddam

## Abstract

The corona virus disease 2019 (COVID-19) is a novel pandemic disease that spreads very fast and causes severe respiratory problem to its carrier and thereby results to death in some cases. In this research, we studied the trend, model Nigeria daily COVID-19 cases and forecast for the future occurrences in the country at large. We adopt the Box and Jenkins approach. The time plot showed that the cases of COVID-19 rises rapidly in recent time. KPSS test confirms the non-stationarity of the process (p < 0.05) before differencing. The test also confirmed the stationarity of the process (p > 0.05) after differencing. Various ARIMA (p,d,q) were examined with their respective AICs and Log-likelihood. ARIMA (1, 2, 1) was selected as the best model due to its least AIC (559.74) and highest log likelihood (−276.87). Both Shapiro-Wilk test and Box test performed confirm the fitness of the model (p > 0.05) for the series. Forecast for 30 days was then made for COVID-19 cases in Nigeria. Conclusively, the model obtained in this research can be used to model, monitor and forecast the daily occurrence of COVID-19 cases in Nigeria.

## 1.0 Introduction

Covid-19 as it is popularly abbreviated means coronavirus disease 2019. This virus surface on the media in December 2019 in a city known as Wuhan is china. It was confirmed to be pandemic according to world health organization WHO (2020a), since the spread of the disease is not only limited to china but to the entire world. The first case of this pandemic outbreak was recorded by the national center for disease control (NCDC, 2020) in Nigeria on 27 February 2020. According to WHO (2020a), The virus was found not to be a living thing but a protein molecule, which makes it difficult to die. The survival of the virus depends on the surface and the environment which determines how they inoculate and incubate. Studies from the literatures show that, the virus which causes severe respiratory problem to its carrier can only disintegrate and decay after it is subjected to high temperature and humidity. Covid-19 can survive in a cold atmosphere because of its adaptation to low temperature, that’s why it spreads faster in countries with lower temperature, and study reveals that countries with higher temperature have lower risk for its spread (WHO, 2020a). Covid-19 needs material medium for its propagation, when it get in contact with the nose, targeted the cells, degraded it and multiply to other part of the respiratory organ including the lungs. When the virus gets contacted with the cells, the body immune system fights back and reduces the efficiency and spread the virus to the entire cell. Due to this, there is high mortality rate in the older people since they have low immune system to fight the virus and this makes the number of recovered cases in the youths to be higher because they have stronger immune system.

According to the world health organization (WHO 2020a), only 2% of the reported cases were below the age of 18 years, and the virus molecule can survive for about 8 hours on paper and fabric while it can survive for more hours on metals, plastic and glass. When an individual has contact with any of these materials which has been contaminated and still in its inoculation period, the virus will temporarily settle on the host, if the host eventually gets to mouth, the eyes or inner part of nose, which are medium to the nasal area, such individual is at high risk on being infected. The initial symptoms include sneezing, itchy eye, dry cough and so on. Recent study shows that, if an individual have closed relationship with a patient with any of the aforementioned symptoms, such individual will be in isolation for at least two weeks, before undergoing a test, if tested positive, will immediately be quarantine for immediate medical care and treatment. In this research, we focus on the study of the trend of the pandemic virus, investigate the causes, develop a suitable model to determine its trend and also use the model to predict future expected developed cases.

## 2.0 Literature Review

Though the COVID-19 is a novel pandemic outbreak, but several authors have made immense contribution towards investigating the outbreak and proffer immediate solution to curb the spread of the disease. Ying et al. (2020) published an up to date review of the literatures on the novel pandemic disease. This shall be discussed in details in this section. According to WHO (2020), the average reproductive number is 1.95, that is, it ranges from 1.4 to 2.5. Joseph et al. (2020) was among recent authors to write about the outbreak of coronavirus in Wuhan, the authors applied the use of the Stochastic Markov chain Monte-Carlo methods with sampling prior using the posterior distribution. The authors discovered that the average reproductive number (R_0_) for the pandemic outbreak is 2.68. i.e. (R_0_=2.68). Shen et al. (2020) developed a model with population divided into five compartments, the authors introduced the non-linear least squares method to get the point estimate with R_0_=6.49, which far greater than the WHO projection. Also, Lin et al (2020) introduce statistical exponential growth with the use of SARS by fitting the exponential growth using poisson regression, the R_0_ was found to be 2.90, which is not too wide than the WHO value. Read et al. (2020) also developed a mathematical transmission model by using the Poisson distribution to assume the daily time increment. The reproductive number was found to be 3.11. The mathematical incidence decay and exponential adjustment model was introduced by Majumder et al. (2020). The authors adopted mean serial interval lengths in fitting the model. Their R_0_ was recorded 2.55, which is very close to the WHO standard.

Recent articles have published in modelling the occurrence and studying the trend of the novel COVID-19 pandemic, such articles include Cori et al (2020), Dung et al (2020), Yuan et al (2020) and WHO (2020b). In this article, instead of using the basic reproductive number in determining the spread of the pandemic outbreak, we intend to adopt a suitable ARIMA model that will assist in forecasting the occurrence of the spread in Nigeria.

## 3.0 Methodology

The Box-Jenkins approach will be adopted in this research and the autoregressive integrated moving average (ARIMA) will be discussed in this section.

### 3.1 Box - Jenkins Approach

The Box-Jenkins refers to a systematic method of identifying, fitting, checking, and using integrated autoregressive, moving average (ARIMA) time series models. The method is appropriate for time series of medium to long length (at least 50 observations). In this research we will adopt the Box-Jenkins method, concentrating on the Arima model. One of the steps in the Box - Jenkins method is to transform a non-stationary series into a stationary one.

### 3.2 Autoregressive Integrated Moving Average Process (ARIMA)

ARIMA models are, in theory, the most general type of models for forecasting a time series which when not stationary can made to be “stationary” by differencing if necessary, probably in conjunction with nonlinear transformations such as logging or deflating. A random variable that is a time series is stationary if its statistical properties are all constant over time. A stationary series has no trend, its variations around its mean have a constant value, and it changes in a consistent way, i.e., its short-term random time patterns always look the same in a statistical sense. The latter condition means that its autocorrelations (correlations with its own prior deviations from the mean) remain constant over time.

There autoregressive moving average (p,q) is of the form

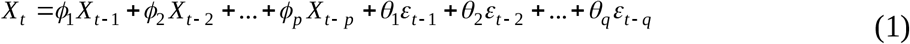

The white noise of the process is expressed as{εt}. In lag form, equation (1) becomes;

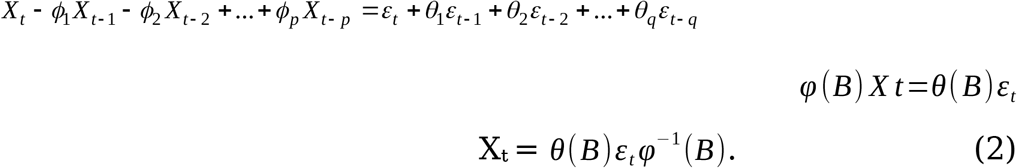

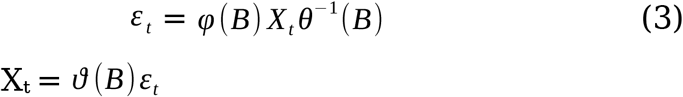

Where

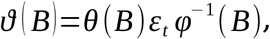

*ϑ* (*B*) is the phi-weight.

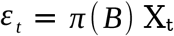

Where,

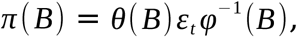

*π* (*B*)represent the pi-weight.

An ARIMA process of order (*p, d, q*) with ARMA (*p, q*) process can be defined by:

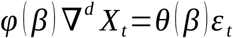

Where p, d, q are non-negative integers.

Note: When d =0, the ARIMA (p, d, q) becomes ARMA(p, q).

In this research the ARIMA model would be applied to the data on Covid-19 to model total daily new cases of the virus from inception February 27 2020 and to further forecast for Total daily occurrence of the virus in the country for a period of time.

### 3.3 Diagnostic checking

The most useful and informative diagnostic checks deal with determining whether the assumptions conditioning the innovation series are satisfied by the residuals of the autoregressive integrated moving average (ARIMA) model. Diagnostic checks only have meaning if the parameters of the model are efficiently estimated using the maximum likelihood approach at the estimation stage. For all of the diagnostic checks presented in this paper, it is assumed that a maximum likelihood estimator is used to estimate the model parameters. The paper discusses Shapiro and box test, whiteness tests, portmanteau tests, normality tests, constant variance tests, and so on.

## 4.0 Analysis and Result

The result of the analysis obtained from daily cases of total Covid-19 in Nigeria between February 27, 2020 and May 1, 2020 is presented in this section. The exploratory analysis, Auto Correlation Function (ACF),Partial Auto Correlation Function (PACF), Augmented Dickey-Fuller (ADF) test, Kwitatkowski Philips Schmidt Shin (KPSS) test, Akaike Information Criterion (AIC) and the Autoregressive Integrated Moving Average (ARIMA) test were carried out, and the result were presented in section 4. More so, the discussion of the analysis is presented in the next section.

### 4.1 Exploratory Data Analysis (EDA) of Covid-19 cases in Nigeria

### 4.2 Time Plots for COVID-19 cases in Nigeria

### 4.3 Unit Root/Stationarity Tests

### 4.4 Autocorrelation Function (ACF) and Partial Autocorrelation Function (PACF)

### 4.4 Autoregressive Integrated Moving Averages (ARIMA) Models Examined

### 4.5 Diagnostic checking

### 4.6 Shapiro - Wilk Test and Box Test

## 5.0 Discussion

Table 1 and Figure 1 show the summary of statistics of Covid-19 daily cases in Nigeria from February 27 to May 1 and the histogram of the distribution of the cases. The exploratory data analysis was done to determine the average case for the time frame used in this research, it was recorded that average daily number of thirty-four (34) cases were reported, with the initial case reported to be 1, this was at the initial stage of the outbreak, and the maximum daily case recorded so far is 238 as at May 1. Both the standard error and variance are very high indicating a high difference among the daily cases. Figures 2 and 3 also show the time plots of total daily Covid-19 cases before and after differencing. The result shows that the process is not stable over time. High and persistence rise is greatly noticed from the plot. Table 2 contains KPSS test statistics. The test was used to confirm the non-stationarity of the process before differencing and stationarity of the process after first differencing.

**Table 1:** Descriptive Statistics.

| Summary of Statistics |  |
| --- | --- |
| Mean | 34 |
| Variance | 3024.05 |
| Standard Deviation | 54.99 |
| Median | 14 |
| Mode | 0 |
| Minimum | 0 |
| Maximum | 238 |

**Table 2:**
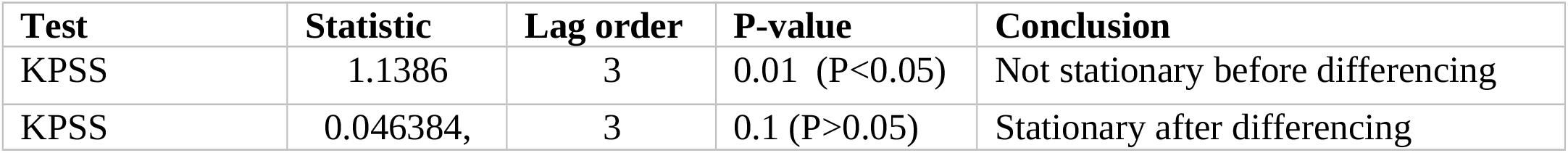
Summary of unit root tests.

**Fig 1:**
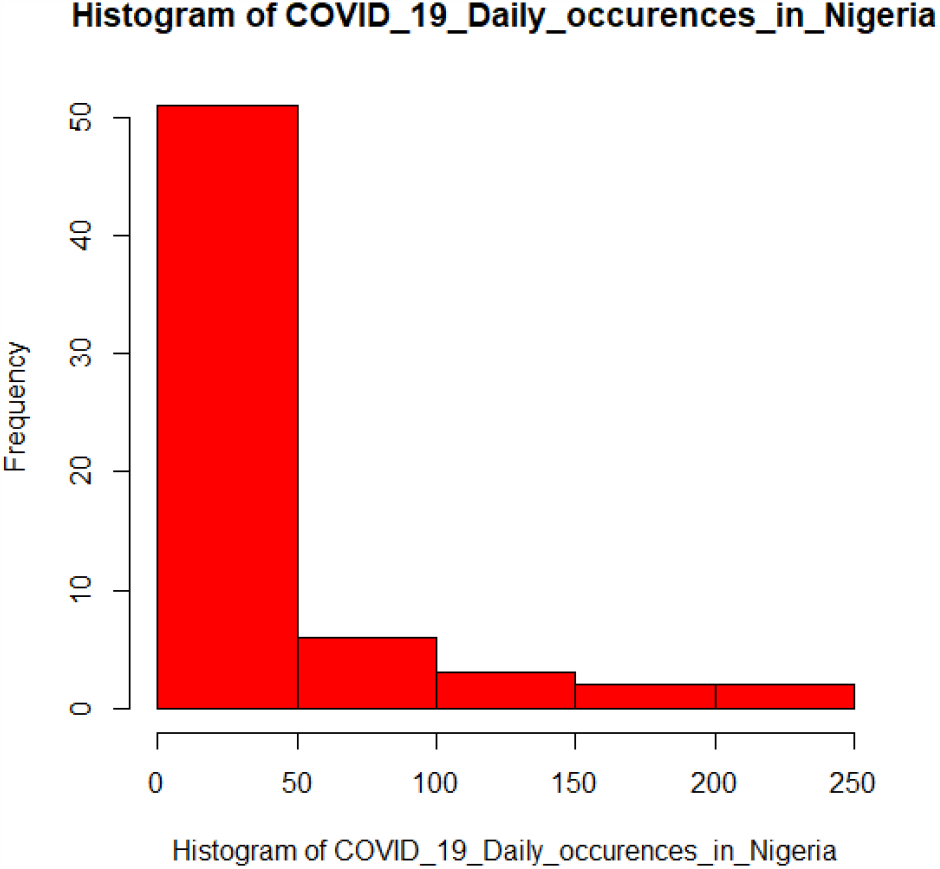
Histogram of Corona daily occurrences.

**Fig 2:**
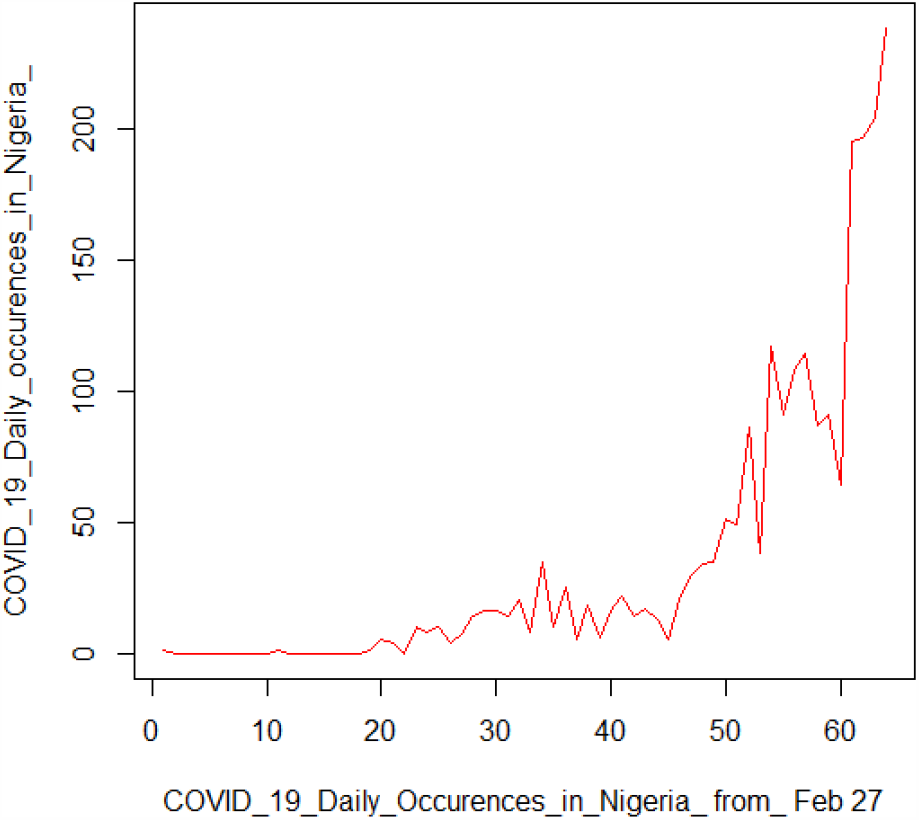
Time plot showing Daily Occurrences of Novel pandemic Covid-19 cases.

**Fig 3:**
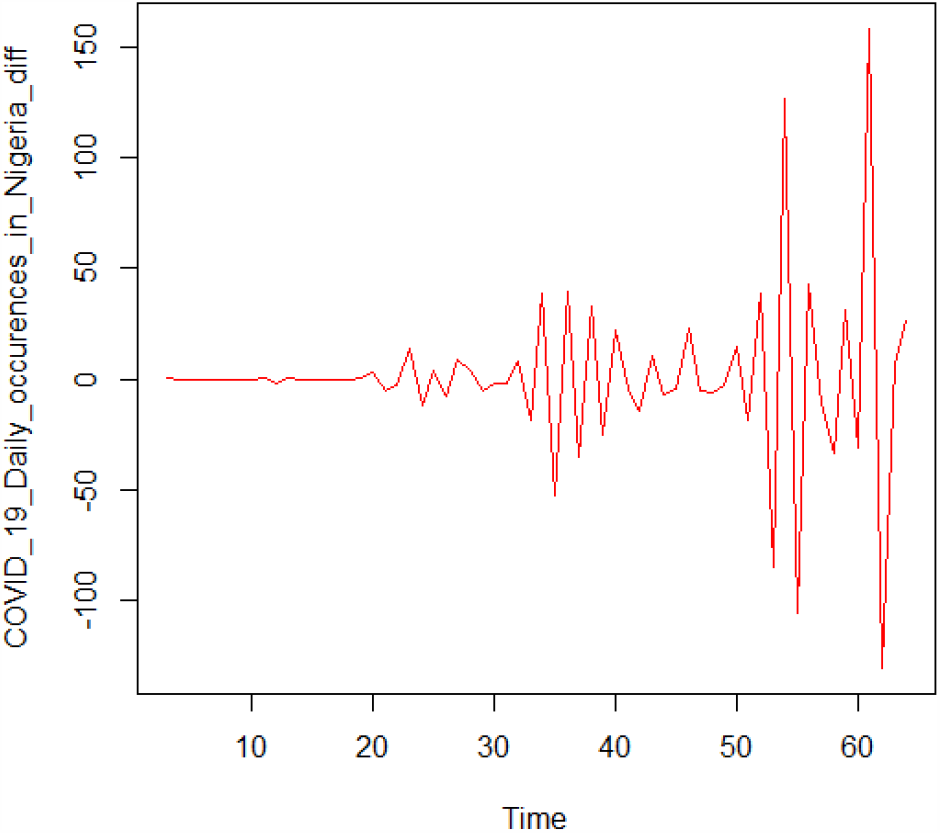
Time plot of the Differenced daily occurrence of Covid-19.

Figures 4 and 5 show the ACF and PACF of the series. The ACF decays slowly while the PACF only exceeds the significant bound at lag 1 and 5 only. Table 3 show the summary of ARIMA (p,d,q) models examined with their AICs and Log-likelihoods. The result showed that ARIMA (1,2,1) has the least AIC (559.74) and the highest Log-likelihood (−276.87). Hence, ARIMA (1,2,1) is selected for the process. The model selected is written as:

**Table 3:** Summary of ARIMA models examined.

| s/n | ARIMA |  |  |  |  |  |  | AIC | Log Likelihood |
| --- | --- | --- | --- | --- | --- | --- | --- | --- | --- |
|  | p | d | q | AR(1) | AR(2) | MA(1) | MA(2) |  |  |
| 1 | 1 | 1 | 0 | -0.4094 | - | - | - | 568.47 | -282.24 |
| 2 | 1 | 1 | 1 | -0.5647 | - | 0.1848 | - | 569.79 | -281.9 |
| 3 | 1 | 1 | 2 | -0.2932 | - | -0.0680 | 0.2179 | 570.43 | -281.22 |
| 4 | 2 | 1 | 2 | -0.3911 | -0.0979 | 0.0258 | 0.2786 | 572.38 | -281.19 |
| 5 | 1 | 2 | 1 | -0.4978 | - | -0.8874 | - | 559.74 | -276.87 |
| 6 | 1 | 2 | 2 | -0.4746 | - | -0.9173 | 0.2691 | 561.73 | -276.86 |

**Fig 4:**
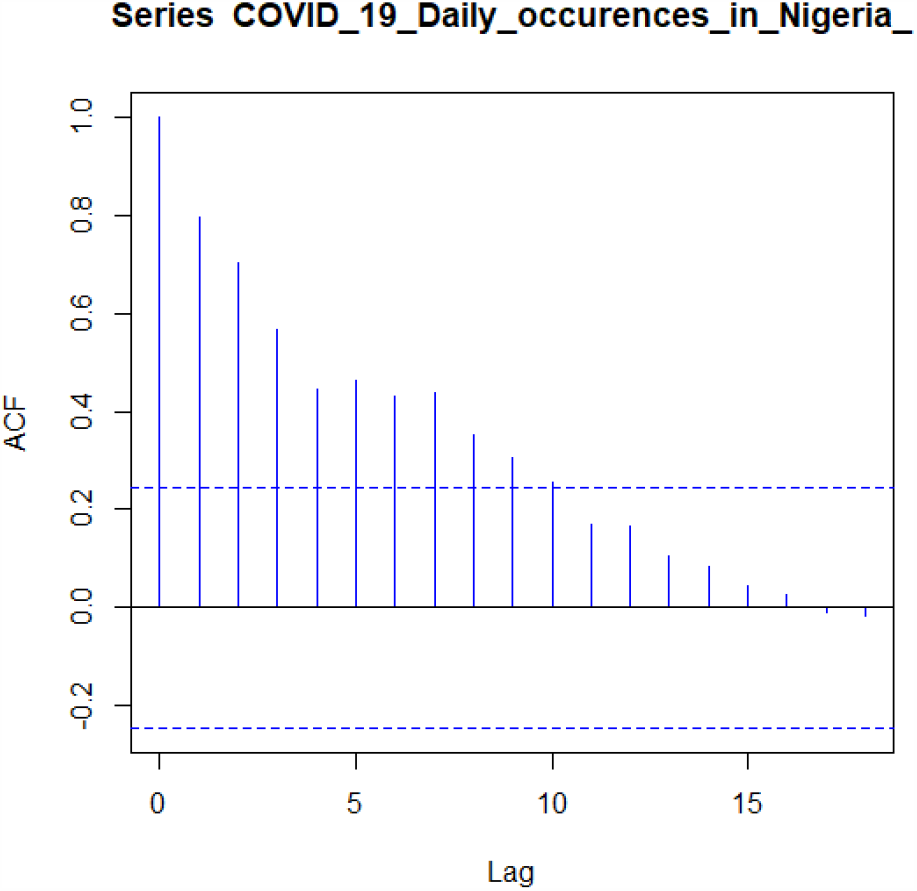
ACF Plot.

**Fig 5:**
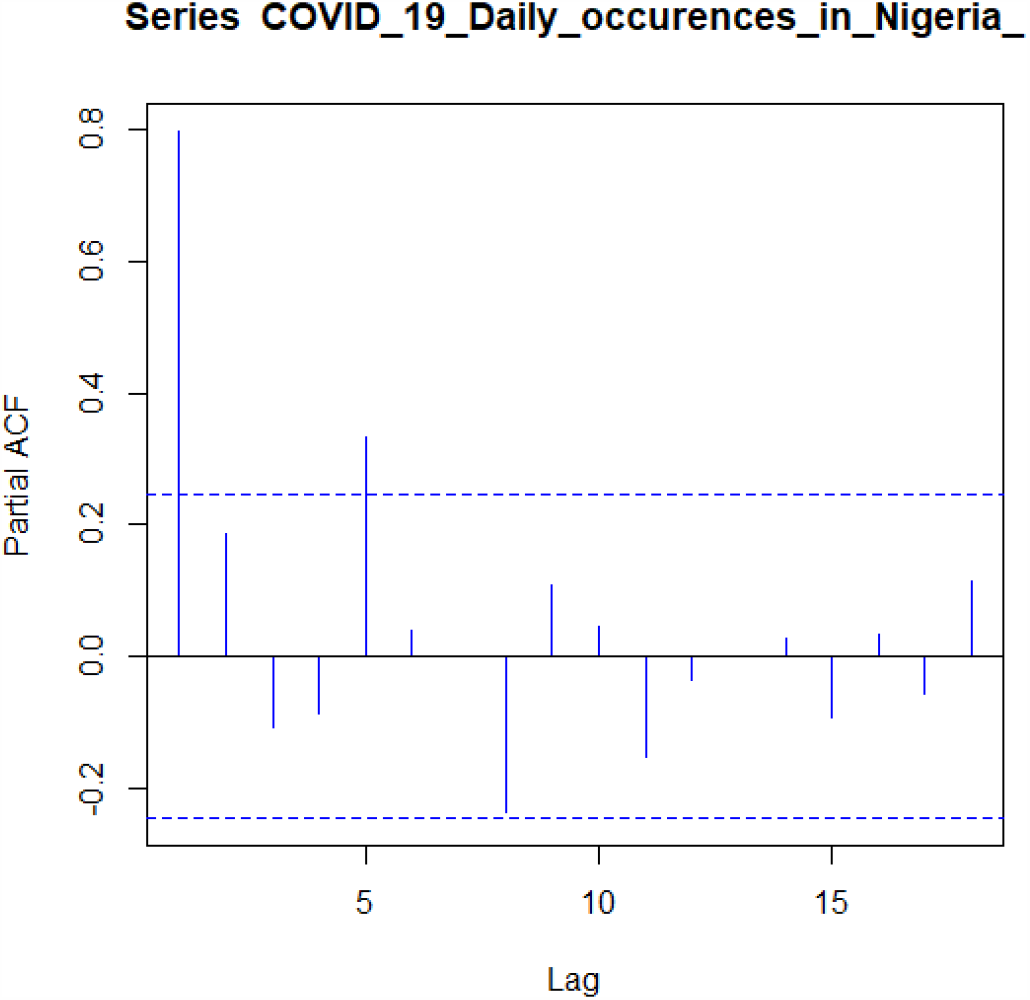
PACF Plot.

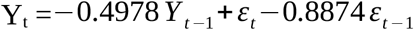

Also, Figure 6 shows the diagnostic plot for the selected model. The results shows that the model fits the process (p-values > 0.5), thereby we fail to reject the null hypothesis and conclude that the model fit the daily occurrence of Covid-19. Table 4 shows Shapiro-Wilk and Box test and the two tests attest to the goodness of fit of the fitted model. Conclusively, forecast for thirty days (May 8, 2020 to June 6, 2020) was then made for the daily cases of COVID-19 cases in Nigeria (see Appendix II) to show how crucial and paramount the need for quick and proactive measure and intervention to curb high and persistence increase in the occurrence of Covid-19 cases.

**Table 4:** Shapiro - Wilk Test and Box Test Results.

| Test | W | P-value | Conclusion |  |
| --- | --- | --- | --- | --- |
| Shapiro-Wilk | 0.67827 | 0.0001 ($P < 0.05$) | The error terms are normally distributed | |
| Test | X-squared | Df | P-Value | Conclusion |
| Box Test | 0.15205 | 1 | 0.6966 ($P > 0.05$) | Residuals are uncorrelated |

**Fig 6:**
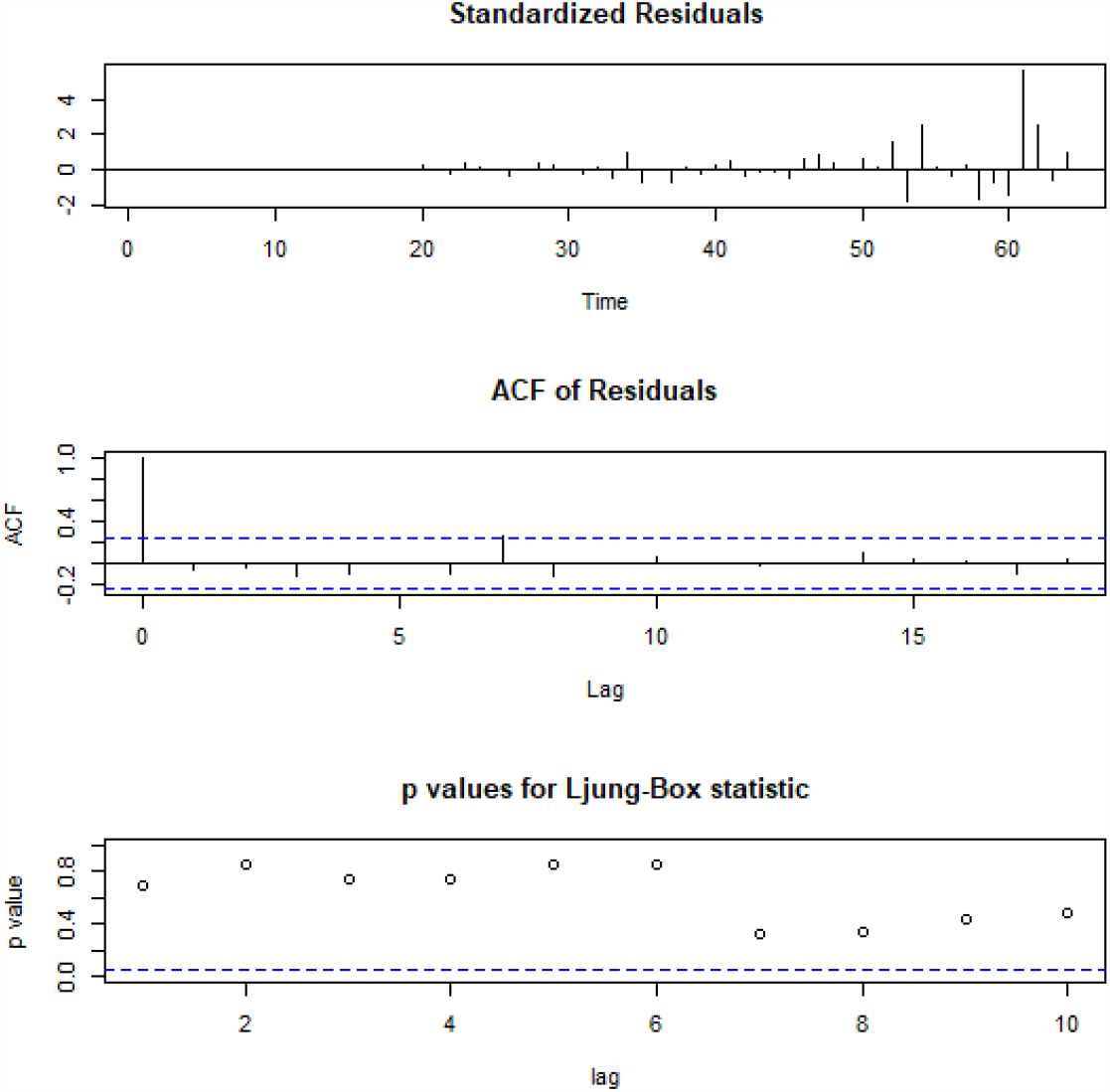
Diagnostic plot.

## 6.0 Conclusion

Succinctly put, the rate of daily occurrence of the novel pandemic Covid-19 outbreak in Nigeria continue to increase every day. Thereby resulting into high loss of lives which may lead to extinction and paralysis of the economy and so its control must be of high concern. Hence, the model obtained from this research can be used to model, monitor and forecast the future case of Covid-19 in Nigeria as a whole so as to put a stop to the high mortality rate caused by the pandemic disease. The fear for the coronavirus is really high among people due to the high rate of its spread and lack of adequate knowledge about the cause(s) and how it spread. Hence, more articles and publications about sensitizing people on the coronavirus need to be made available. Also, preventive measure highlighted by WHO such as sanitization of hand regularly, avoiding direct contact with eyes, mouth and nose with unclean hands, keeping distance of at least one meter, avoiding handshake, not travelling while one is sick and seeking medical attention when symptoms of the Covid-19 such as high fever and so on are observed are hereby recommended as solutions to avoid quick spread of the diseases. It is expected that effective implementation of the above recommendations will guide and help in preventing the spread of the disease and thereby reduces its daily occurrences.

## Data Availability

The data is available at the National centre for disease control website

## Appendix II

**Table 5:** Covid-19 daily occurrence from Feb 27, 2020 to May 1, 2020.

| DAILY OCCURENCES | CONFIRMED CASES | NEW CASES | DISCHARGED | DEATHS |
| --- | --- | --- | --- | --- |
| 2/27/2020 | 1 | 1 | 0 | 0 |
| 2/28/2020 | 0 | 0 | 0 | 0 |
| 3/1/2020 | 0 | 0 | 0 | 0 |
| 3/2/2020 | 0 | 0 | 0 | 0 |
| 3/3/2020 | 0 | 0 | 0 | 0 |
| 3/4/2020 | 0 | 0 | 0 | 0 |
| 3/5/2020 | 0 | 0 | 0 | 0 |
| 3/6/2020 | 0 | 0 | 0 | 0 |
| 3/7/2020 | 0 | 0 | 0 | 0 |
| 3/8/2020 | 0 | 0 | 0 | 0 |
| 3/9/2020 | 2 | 1 | 0 | 0 |
| 3/10/2020 | 0 | 0 | 0 | 0 |
| 3/11/2020 | 0 | 0 | 0 | 0 |
| 3/12/2020 | 0 | 0 | 0 | 0 |
| 3/13/2020 | 0 | 0 | 0 | 0 |
| 3/14/2020 | 0 | 0 | 0 | 0 |
| 3/15/2020 | 0 | 0 | 0 | 0 |
| 3/16/2020 | 0 | 0 | 0 | 0 |
| 3/17/2020 | 3 | 1 | 0 | 0 |
| 3/18/2020 | 8 | 5 | 0 | 0 |
| 3/19/2020 | 12 | 4 | 0 | 0 |
| 3/20/2020 | 12 | 0 | 0 | 0 |
| 3/21/2020 | 22 | 10 | 2 | 0 |
| 3/22/2020 | 30 | 8 | 0 | 0 |
| 3/23/2020 | 40 | 10 | 0 | 0 |
| 3/24/2020 | 44 | 4 | 0 | 1 |
| 3/25/2020 | 51 | 7 | 0 | 0 |
| 3/26/2020 | 65 | 14 | 1 | 0 |
| 3/27/2020 | 81 | 16 | 0 | 0 |
| 3/28/2020 | 97 | 16 | 0 | 0 |
| 3/29/2020 | 111 | 14 | 0 | 0 |
| 3/30/2020 | 131 | 20 | 5 | 0 |
| 3/31/2020 | 139 | 8 | 1 | 1 |
| 4/1/2020 | 174 | 35 | 0 | 0 |
| 4/2/2020 | 184 | 10 | 11 | 0 |
| 4/3/2020 | 209 | 25 | 5 | 2 |
| 4/4/2020 | 214 | 5 | 0 | 0 |
| 4/5/2020 | 232 | 18 | 8 | 1 |
| 4/6/2020 | 238 | 6 | 2 | 0 |
| 4/7/2020 | 254 | 16 | 9 | 1 |
| 4/8/2020 | 276 | 22 | 0 | 0 |
| 4/9/2020 | 288 | 14 | 7 | 1 |
| 4/10/2020 | 305 | 17 | 7 | 0 |
| 4/11/2020 | 318 | 13 | 12 | 3 |
| 4/12/2020 | 323 | 5 | 15 | 0 |
| 4/13/2020 | 343 | 20 | 6 | 0 |
| 4/14/2020 | 373 | 30 | 8 | 1 |
| 4/15/2020 | 407 | 34 | 29 | 1 |
| 4/16/2020 | 442 | 35 | 24 | 1 |
| 4/17/2020 | 493 | 51 | 7 | 4 |
| 4/18/2020 | 542 | 49 | 7 | 2 |
| 4/19/2020 | 627 | 86 | 4 | 2 |
| 4/20/2020 | 665 | 38 | 18 | 1 |
| 4/21/2020 | 782 | 117 | 9 | 3 |
| 4/22/2020 | 873 | 91 | 0 | 3 |
| 4/23/2020 | 981 | 108 | 0 | 3 |
| 4/24/2020 | 1095 | 114 | 11 | 1 |
| 4/25/2020 | 1182 | 87 | 14 | 3 |
| 4/26/2020 | 1273 | 91 | 17 | 5 |
| 4/27/2020 | 1337 | 64 | 16 | 0 |
| 4/28/2020 | 1532 | 195 | 4 | 4 |
| 4/29/2020 | 1728 | 196 | 52 | 7 |
| 4/30/2020 | 1932 | 204 | 12 | 7 |
| 5/1/2020 | 2170 | 238 | 32 | 10 |

**\*Source:** www.covid19.ncdc.gov.ng

**Table 6:**
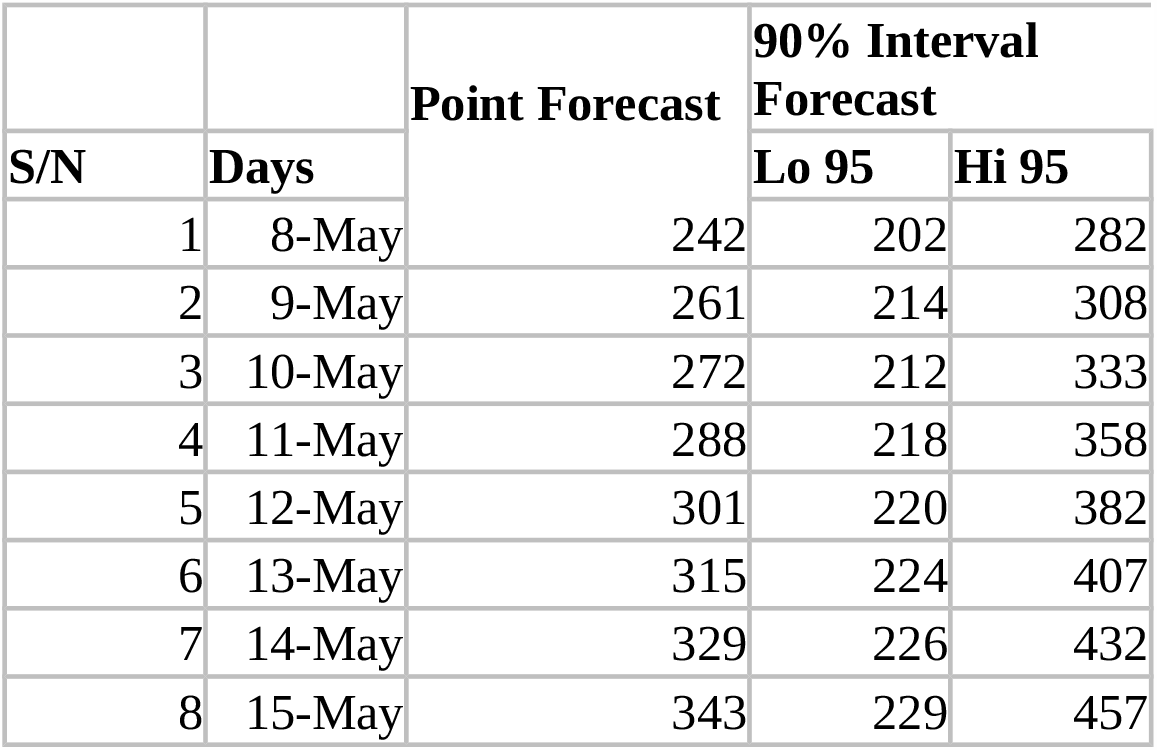
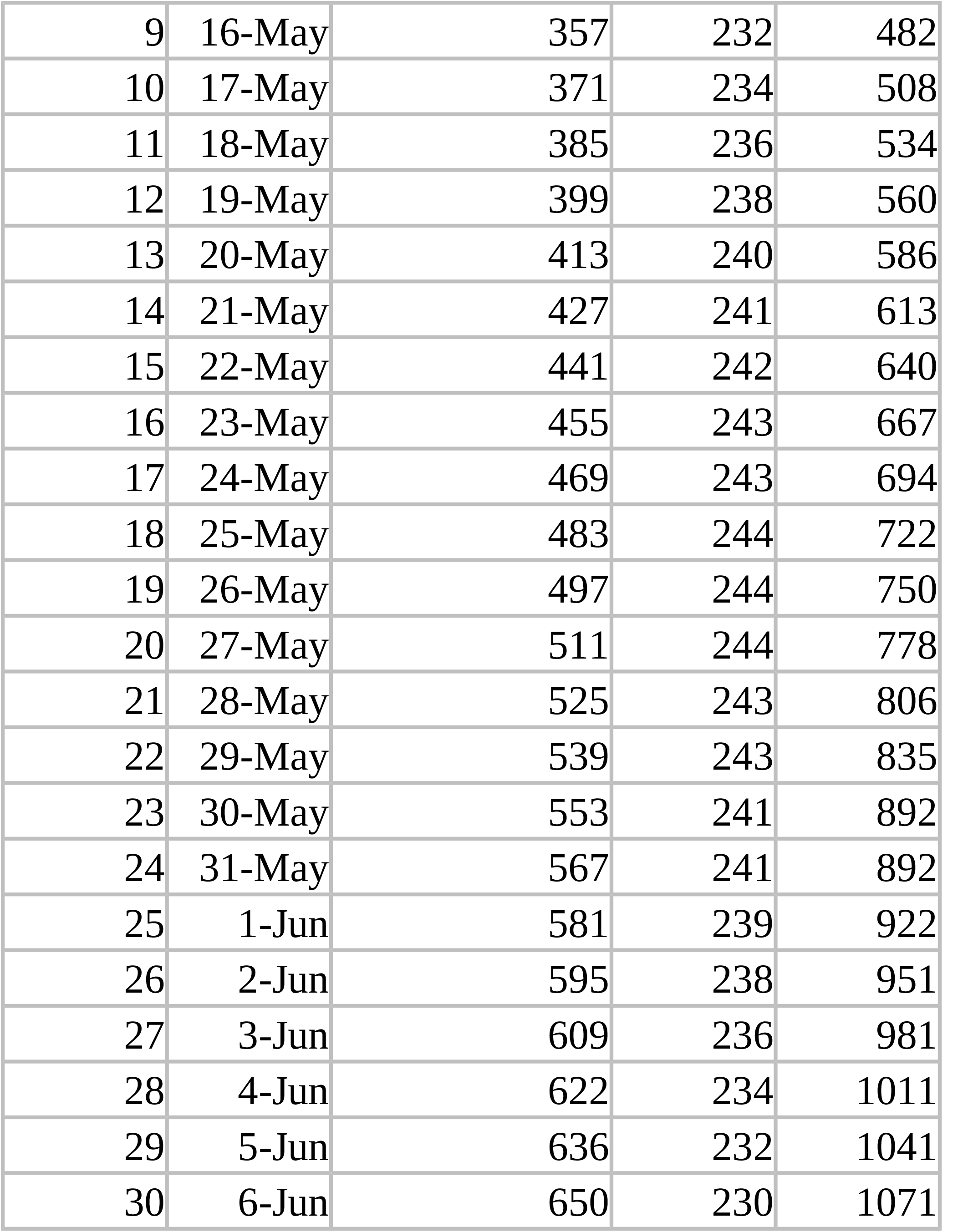
Thirty Days point and interval forecast for Covid-19 cases in Nigeria from May 8 – June 6.

| S/N | Days | Point Forecast | 90% Interval Forecast |  |
| --- | --- | --- | --- | --- |
|  |  |  | Lo 95 | Hi 95 |
| 1 | 8-May | 242 | 202 | 282 |
| 2 | 9-May | 261 | 214 | 308 |
| 3 | 10-May | 272 | 212 | 333 |
| 4 | 11-May | 288 | 218 | 358 |
| 5 | 12-May | 301 | 220 | 382 |
| 6 | 13-May | 315 | 224 | 407 |
| 7 | 14-May | 329 | 226 | 432 |
| 8 | 15-May | 343 | 229 | 457 |
| 9 | 16-May | 357 | 232 | 482 |
| 10 | 17-May | 371 | 234 | 508 |
| 11 | 18-May | 385 | 236 | 534 |
| 12 | 19-May | 399 | 238 | 560 |
| 13 | 20-May | 413 | 240 | 586 |
| 14 | 21-May | 427 | 241 | 613 |
| 15 | 22-May | 441 | 242 | 640 |
| 16 | 23-May | 455 | 243 | 667 |
| 17 | 24-May | 469 | 243 | 694 |
| 18 | 25-May | 483 | 244 | 722 |
| 19 | 26-May | 497 | 244 | 750 |
| 20 | 27-May | 511 | 244 | 778 |
| 21 | 28-May | 525 | 243 | 806 |
| 22 | 29-May | 539 | 243 | 835 |
| 23 | 30-May | 553 | 241 | 892 |
| 24 | 31-May | 567 | 241 | 892 |
| 25 | 1-Jun | 581 | 239 | 922 |
| 26 | 2-Jun | 595 | 238 | 951 |
| 27 | 3-Jun | 609 | 236 | 981 |
| 28 | 4-Jun | 622 | 234 | 1011 |
| 29 | 5-Jun | 636 | 232 | 1041 |
| 30 | 6-Jun | 650 | 230 | 1071 |

